## Supplementary Materials for "Age-seroprevalence curves for the multi-strain structure of influenza A virus"

### 1 Validation of PC1 as a surrogate for 'composite seroprevalence'

To validate the first principal component as a viable surrogate for the seroprevalence, we built an individual-based simulation of influenza antibody acquisition and waning – as it would be expected to occur in Vietnam and with no influenza vaccination. The simulation was built in C++ and the source code is available at https://github.com/bonilab/seroepi-02FL-influenza-vietnam-PCA.

#### *1.1 Challenges*

There are several challenges in developing an individual-based epidemiological simulation for seasonal influenza. Controlling the amount of infection and the seasonality of infection in an individual-based model (IBM) is straightforward, as we can either vary the transmission parameter or fix the number of annual infections and distribute them according to a pre-defined season or epidemic timing. Modeling the antibody titer post-infection and the waning of antibody post-infection is straightforward as these have both been estimated from large observational studies (Todd 2016). Likewise, the protective effect of an antibody titer is known, and the protective effect of protein microarray (PMA) antibody titers was measured in a case-control study run in parallel with this sample collection (Todd 2016).

However, although we know the protective effect of antibody titer against becoming an *influenza case*, we do not know the protective effect against becoming an *influenza infection*. Although older studies have measured protection against infection with a household design, the sample sizes in these studies were generally small. For our PMA assay, this type of study was not carried out, and all we know is the degree to which a titer protects someone from becoming a symptomatic *case* of influenza (Figure S1).

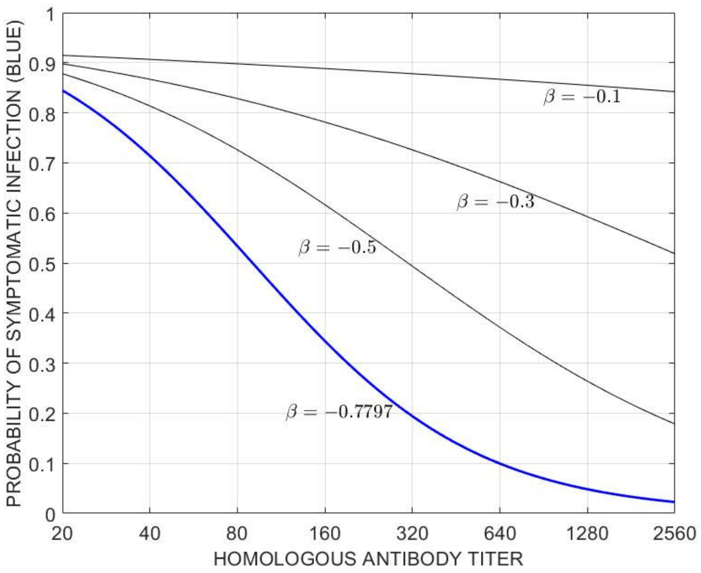


**Figure S1**. Blue line shows probability of symptomatic infection from a test-negative case-control design (N=814) from a logistic regression. Equation is $prob = c\, e^{-\beta \cdot titer} / (1 + c\, e^{-\beta \cdot titer})$, with $c$ = 11.87 and $\beta$ = -0.7797 as the inferred values. Curves for three other $\beta$-values are shown for the likely relationship between titer and protection from influenza infection (symptomatic or asymptomatic).

To verify that infections occur at the right frequency in the correct titer groups in our simulation, we could (as a proxy) ensure that infections occur at the right frequency to the right age groups. But, the *age-distribution of infections* in an influenza epidemic is not known – because (*i*) it is different in every epidemic and (*ii*) age-stratified serology studies captures the age distribution of all past infections but not the age distribution of infections in any particular epidemic or year.

Our hunch is that $\beta$=-0.3 is about the right relationship for Figure S1's protective effect against infection. Titers of 200 and 300 are very common in the data set and it is unlikely that these individuals would be highly protected from infection (making

$\beta$=-0.5 less likely). Individuals with very high titers (~1000 or ~2000, approaching post-infection titers) are probably protected from infection (however, there is no data on this) making $\beta$=-0.1 an unlikely parameter value.

Second, cross-reactivity among strains determines the interpretation of the PCA. If there is no cross-reactivity, PC1 counts the total number of past infections. If there is complete cross-reactivity, PC1 should track whether a person has ever been infected. A mathematical relationship between the cross-reactivity parameter $\sigma$ and PC1 is needed to describe this quantity of past exposure and its epidemiological interpretation. For the present work, we measure the Pearson correlation between neighboring strains (i.e. strains that are two years apart in our strain set for H3) and average it across the four neighbor-pairs to obtain $\sigma = 0.88345$, and we use $\sigma = 0.88$ for the simulation runs below. In other words, an infection with H3-2007 generating an H3-2007 antibody titer of 1000 would generate antibody titers of ~880 to H3-2005 and H3-2009. For other pathogens, the $\sigma$-values will differ and the PC1 interpretations will differ. Because $\sigma$ is close to one for influenza H3, the PC1 as seroprevalence relationship is the appropriate one to investigate.

Third, in order to mimic the epidemiological characteristics of southern Vietnam, we use influenza case data showing the timing and seasonality (or lack thereof) of influenza epidemics over the past ten years, and we fix the attack rate (which was not measured) in these data by inflating/deflating case numbers by appropriate amounts. However, with a fixed attack rate and fixed season every year, a simulation like this lacks flexibility, and in order to 'meet' high attack-rate requirements during some periods, the simulation may over-infect children as these are the individuals with the lowest antibody titers in the simulation. This means that for higher fixed attack rates the number of infections in children may not be reliable, and this is important because the seroprevalence in children is the primary determinant of long-term average annual attack rate. We can tell that simulation behaviors like this occur at high attack rates because of the high variance in simulation outcomes when attack rates are fixed to be high (together with inspection of results from individual simulation runs).

#### *1.2 Simulation Details*

First, we do not develop a traditional *mass-action* simulation. Rather, we develop a *fixed attack-rate* simulation in order to mimic Vietnam's pattern of annual infections, without modeling the explicit infection process based on number of available susceptibles. A traditional *mass-action* simulation is the right mechanistic approach for studying influenza dynamics, but we are not guaranteed to be able to recover ten years of Vietnam's influenza dynamics with this simulation approach.

An individual-based simulation was developed in C++ to mimic the time pattern and age-pattern of influenza A/H3N2 infections in Vietnam. Using data from Vietnam's National Influenza Surveillance System (Nguyen et al 2009, 2013; Pham et al 2014), we assembled a 9-year time series (2006-2014) of weekly influenza A/H3N2 infections, scaled using the reported number of influenza-like illness (ILI) presentations; in other words, this is the weekly ILI+ time series for H3N2. We concatenated the time series together four times to generate a 36-year ILI+ trend, and simulated weekly infections in an *in silico* population (500,000 individuals) by infecting a new number of individuals every week according the number of cases designated for that week. During weeks with large numbers of infections, it appeared that the simulation infected more children than usual, but we have no way of verifying whether this is supported by data (remember, these are *infections* not *cases*). The concatenated time series was scaled for a 25% annual attack rate, and the time series was inflated/deflated by a multiplicative factor to investigate attack rates between 5% and 30%.

The simulation keeps track of 18 hypothetical flu antigens (strains) each of which circulates for exactly two years. Individuals infected in a given two-year period are infected with that strain. Each individual has 18 antibody titers to these strains that start out at a value 10 (the detection limit) at birth, and the individual acquires higher titers to strains and neighboring strains upon infection. Individuals are chosen for infection every week according to the probabilities shown in Figure S1, and if not enough individuals are chosen the algorithm will continue attempting to infect individuals until the right amount for that week have been infected. Once an individual is infected a titer, a normally distributed log2-titer (mean=7.72, sd=1.311, see Todd (2016)) is drawn. Neighboring strains two years away are given a titer that is smaller by a factor $\sigma$. Neighboring strains four years away are given a titer that is smaller by a factor $\sigma^2$. Neighboring strains six years away are given a titer that is smaller by a factor

$\sigma^3$ (while always ensuring that titers are not lowered after an infection). Antibody waning occurs at a rate of 2.86-log2 units per year (Todd 2016), and waning stops after one year (this is unknown).

An additional feature of back-boosting was added to the simulation (Fonville et al, 2014). When an individual was infected, all past titers that had some signal of past positivity (arbitrarily chosen at titer>50) were boosted by a factor of 7.26 (=$2^{2.86}$) so that they would wane back down to their previous levels after one year.

Sampling was conducted at exactly the same times as in our study samples. 6700 samples were collected in the simulation to mimic sample collection in Ho Chi Minh City, and a population of 500,000 individuals was simulated, as simulations larger than this did not have more stability (tested through trial and error).

Age-structure and age-specific birth and death rate for Vietnam were obtained from the General Statistics Office of Vietnam (https://www.gso.gov.vn/en/data-and-statistics/2019/05/the-1-4-2014-viet-nam-intercensal-population-and-housing-survey-major-findings/).

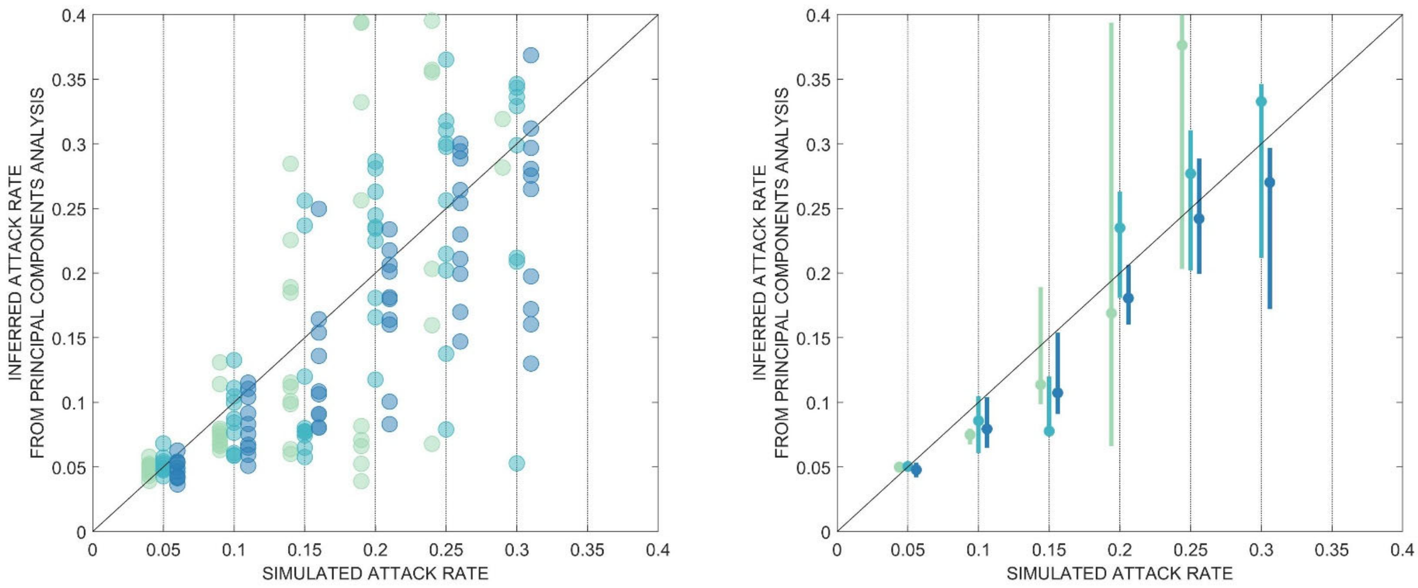


**Figure S2.** Comparison of inferred attack rates from principal components analysis and fixed attack rates in simulation. An empirically measured cross-reactivity parameter $\sigma$ = 0.88 was used for neighboring strains. In both panels, the three different color shades, from left to right, represent $\beta$ = -0.5 (mint green), $\beta$ = -0.3 (sea green), and $\beta$ = -0.1 (dark blue). The left panel plots all ten replicates for each combination of attack rate and $\beta$. Right panel shows the exact same simulation output but with medians and interquartile bars. Fixed attack rates are always in multiples of five, and points and lines are offset horizontally for improved clarity.

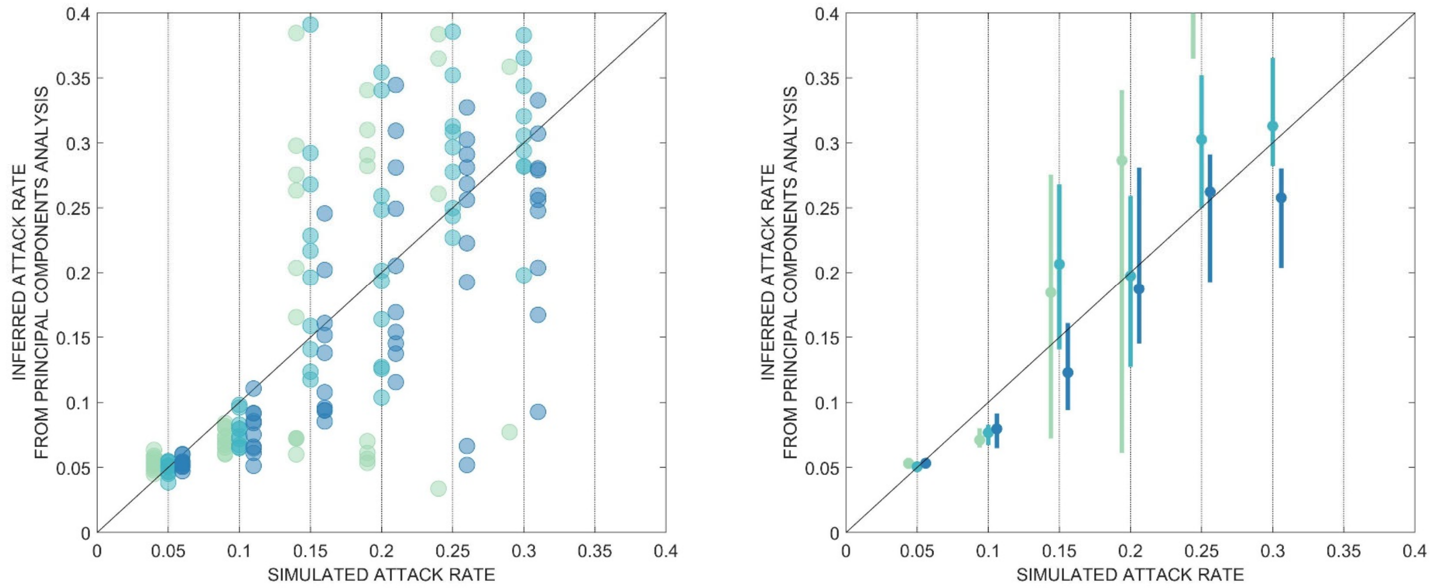


**Figure S3.** Comparison of inferred attack rates from principal components analysis and fixed attack rates in simulation, <u>when back-boosting as described by Fonville et al (2014) is included</u>. The titer of an individual's first infection is set to 1.5 the typical amount (typical=2108, 1.5x=3162), and titers for all neighboring strains are affected accordingly. An empirically measured cross-reactivity parameter $\sigma = 0.88$ was used for neighboring strains. In both panels, the three different color shades, from left to right, represent $\beta$ = -0.5 (mint green), $\beta$ = -0.3 (sea green), and $\beta$ = -0.1 (dark blue). The left panel plots all ten replicates for each combination of attack rate and $\beta$. Right panel shows the exact same simulation output but with medians and interquartile bars. Fixed attack rates are always in multiples of five, and points and lines are offset horizontally for improved clarity.

Figures S2 and S3 show that the PC1-inferred attack rate is a reasonable surrogate for the simulated attack rate, except for attack rates above 25% when $\beta$ = -0.5. The estimates are also biased downwards when the attack rate is 10%. The real concern for this type of validation exercise is the amount of variation in the inferred attack rates when the simulated attack rate is high, which results from the simulation's high variance in levels of infection in children, which may of course be a real phenomenon for influenza epidemics. The next steps in refining PCA as a useful tool for seroprevalence estimation in antigenically variable pathogens are (1) determining the nature of the mathematical relationships among PC1, $\sigma$, and the probability of past infection, and (2) understanding if this relationship can be used to correctly infer long-term attack rates even when infection rates in children fluctuate from year to year with high variance.

### 2 Out-of-sample cross-validation on PC basis change

To perform an internal cross-validation of the stability of the PCA basis change, we broke the data into five subsets ($n$=4880 samples each, randomly assigned) and carried out a PCA on each subset to determine the basis change from antigenic coordinates to principal coordinates. Visually, the five basis change matrices looked very similar. Mapping individuals in subset $j$ with a basis change determined by subset $k$ did not have a substantial effect on the attack rate inference. However, the subsets themselves had different inferred attack rates, as the attack rate inference is sensitive to the number of children under five in each subset.

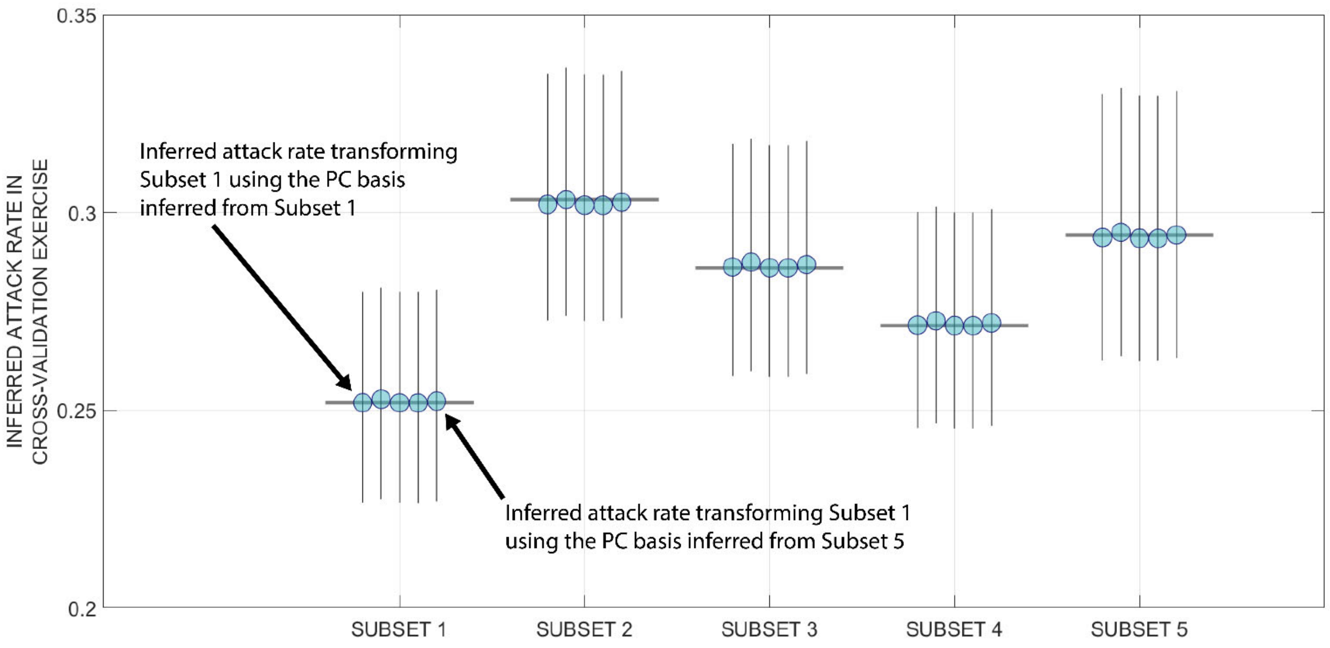


**Figure S4.** Inferred attack rate for five different subsets of the data. Subsets 1 through 4 have 4880 samples and subset 5 has 4882 samples. For each data subset, we performed five different basis changes from antigenic coordinates to principal coordinates, and these are the five circles shown for each subset (in order, from left to right). For each basis change on each subset, we constructed a 4880 × 2 matrix showing how PC1 changes with age and this PC1-age relationship was used to infer the attack rate. 95% confidence intervals were obtained through likelihood profiling and are shown as vertical black lines. The structure of principal components space is fairly stable and does not change when we look at subsets of the data. However, subsets of the data will vary in the number of samples available from young children, and for this reason the attack rate estimates vary from 25% to 30% across the different subsets.

Supplementary Appendix for "Age-seroprevalence curves for the multi-strain structure of influenza A virus"

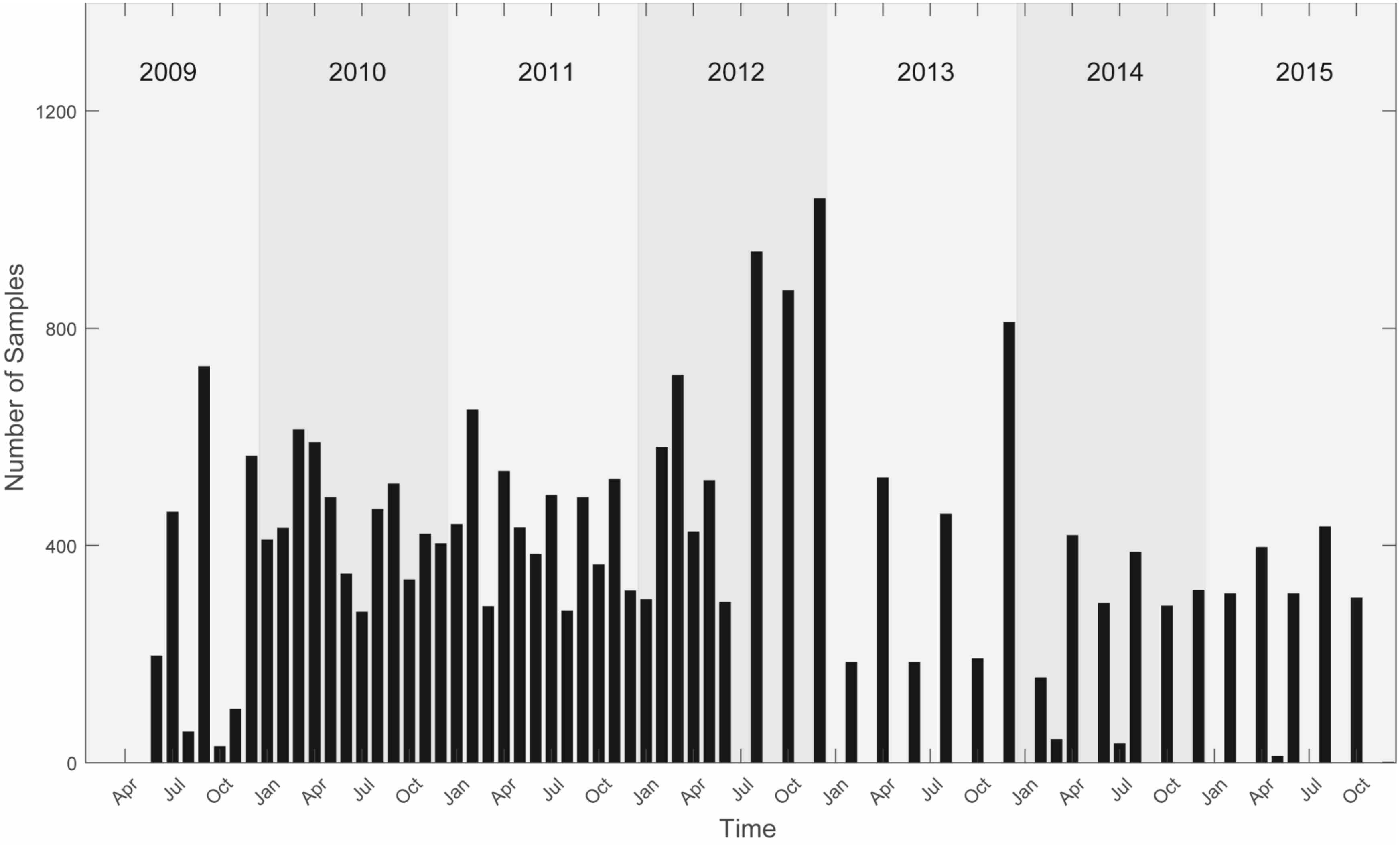


**Figure S5.** Collection times of the 24402 serum samples in the study. The black bars show the number of serum samples collected from all sites in the corresponding month. Two consecutive years are highlighted with different gray scales.

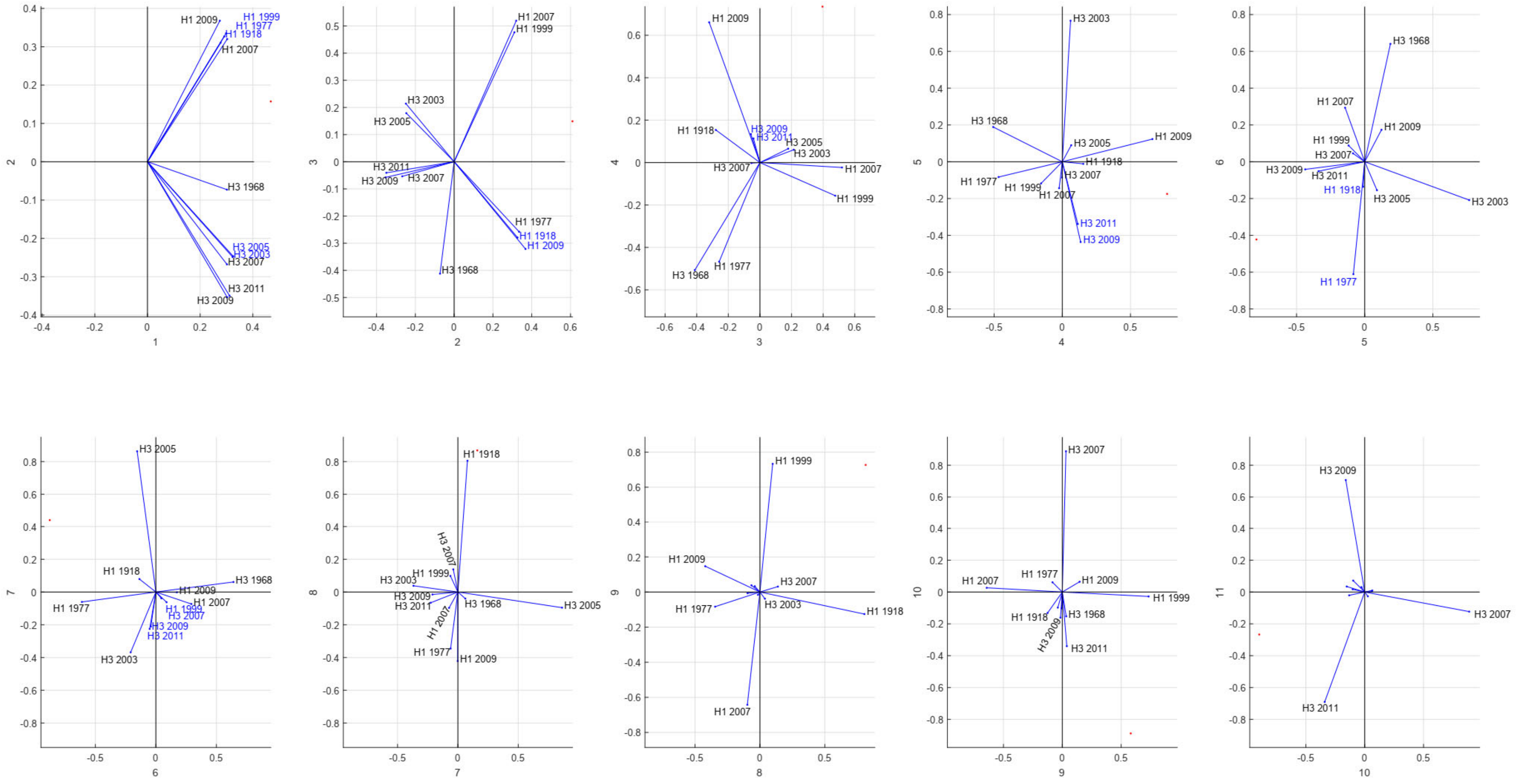


**Figure S6.** Principal component (PC) loadings for 11 different antigens (six H3N2 and five H1N1 trains) together. Only two consecutive components are shown in each panel. Antigen names whose coordinates are almost identical in the given PC-space are in blue.

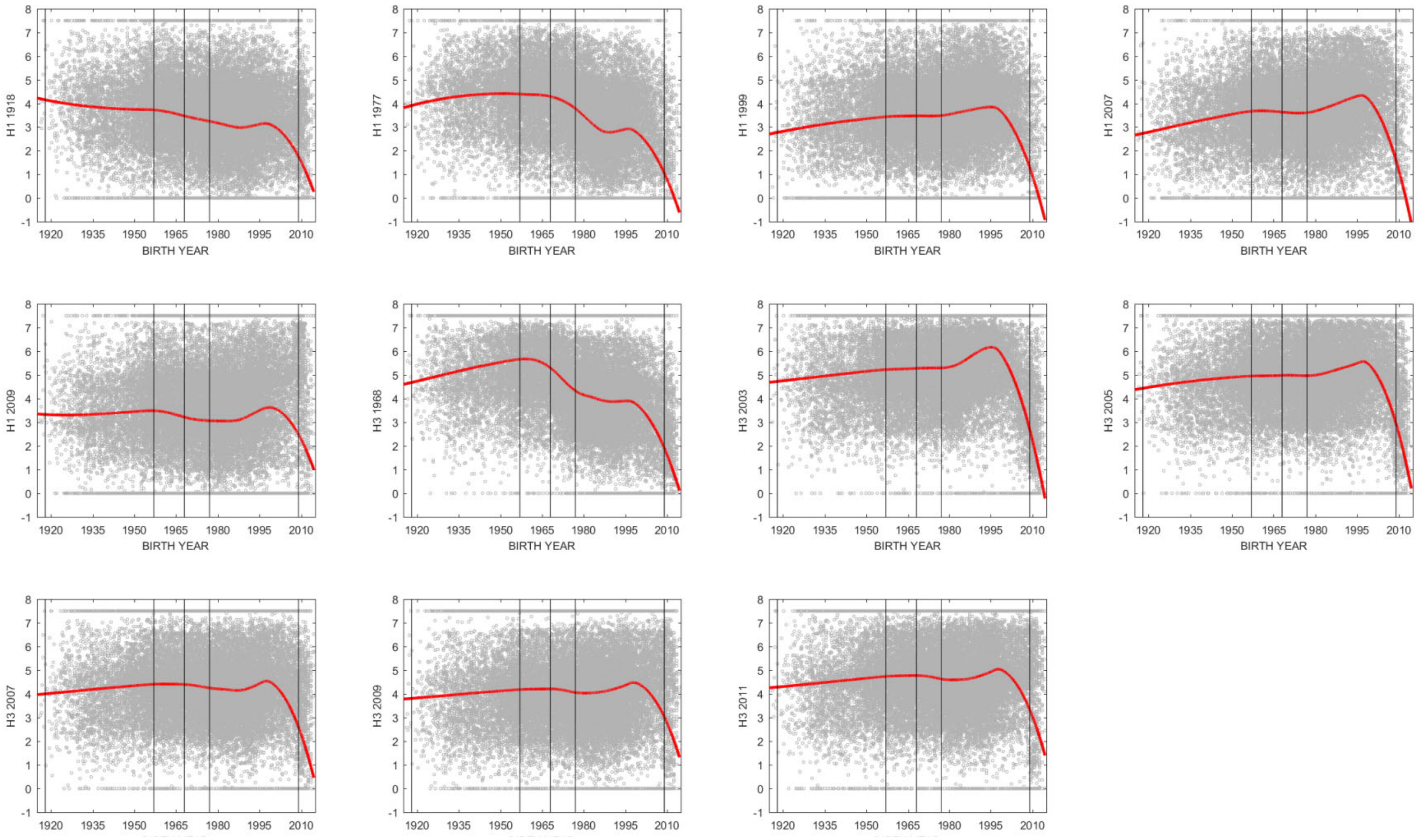


**Figure S7.** Relationship between birth year and antibody titers of six H3N2 and five H1N1 strains. Each panel corresponds to an influenza antigen. Gray dots show log-titer (*y*-axis) for individuals in the data set. Red curves are LOESS regressions (spanning factor=0.50. Vertical black lines mark the introductions of new influenza subtypes. Here, the smoothed red curves look like age-seroprevalence curves as antibody titers generally increase with age.

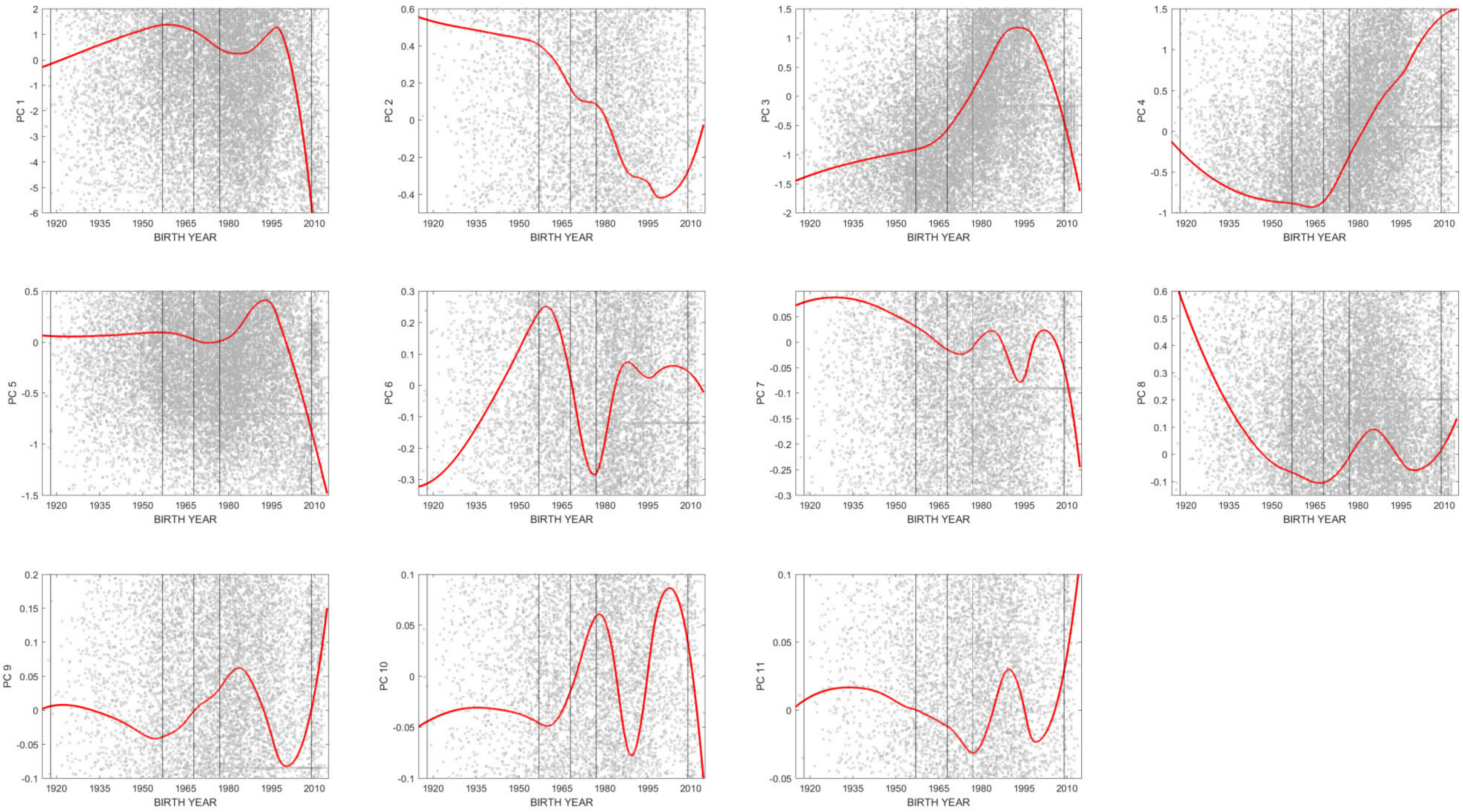


**Figure S8.** Relationship between birth year and the eleven principal components. Gray dots show log-titer (*y*-axis) for individuals in the data set. Red curves are LOESS regressions (spanning factor=0.50). Vertical black lines mark the introductions of new influenza subtypes. PC2 distinguishes individuals who have stronger H1 responses (high values) and stronger H3 responses (low values). PC3 distinguishes individuals by their H1 exposure, whether primary exposure was to the interpandemic H1N1 lineages (1999 and 2007 strains, high values) or to 2009 H1N1-like or 1918-like lineages (low values). PC4 distinguishes the 2009 and 1918 H1N1 viruses (high values) from the 1977 pandemic H1N1 virus (low values). Other PC-based distinctions can be inferred using the loadings in Figure S2.

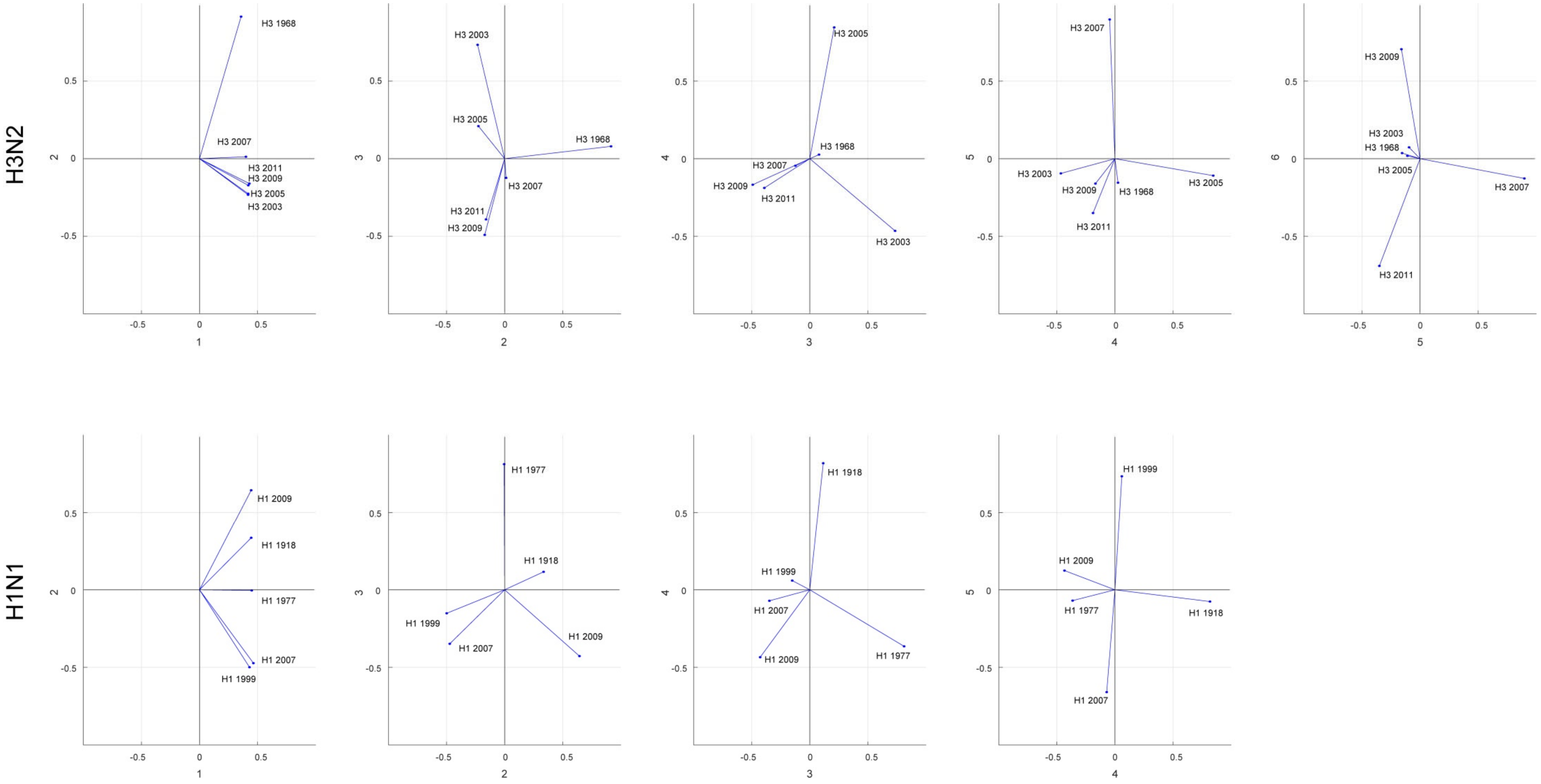


**Figure S9.** Principal component loadings for H3N2 antigens and H1N1 antigens, with PCAs done separate for the two antigens.

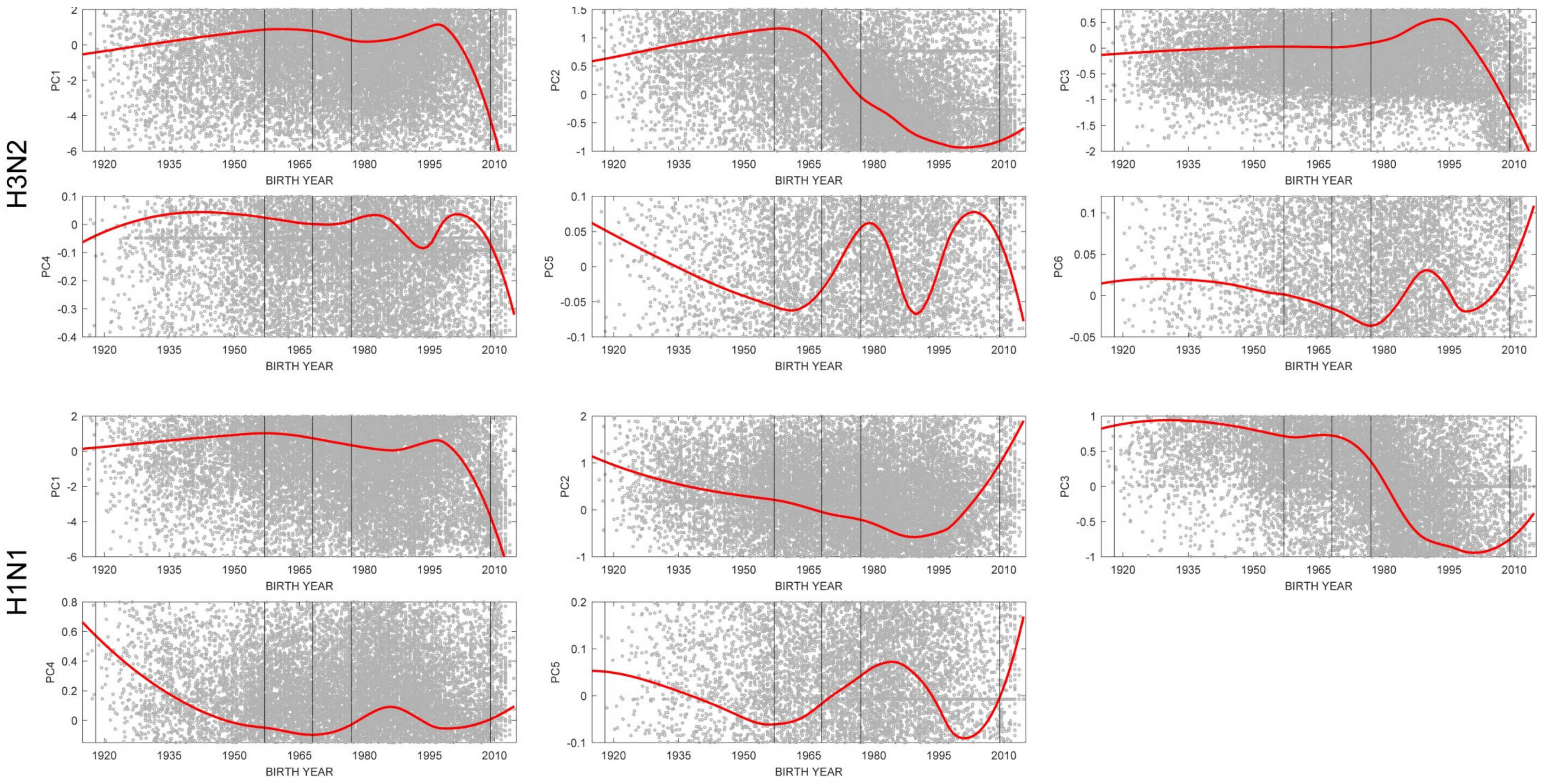


**Figure S10.** Relationship between birth year and six H3N2 PCs and five H1N1 PCs that are presented in Figure S5. For H3N2 viruses, PC2 distinguishes the 1968 virus from the 2003-2011 H3N2 strains. For H1N1, PC2 distinguishes the inter-pandemic strains (1999 and 2007) from the 1918/2009 antigenic types.

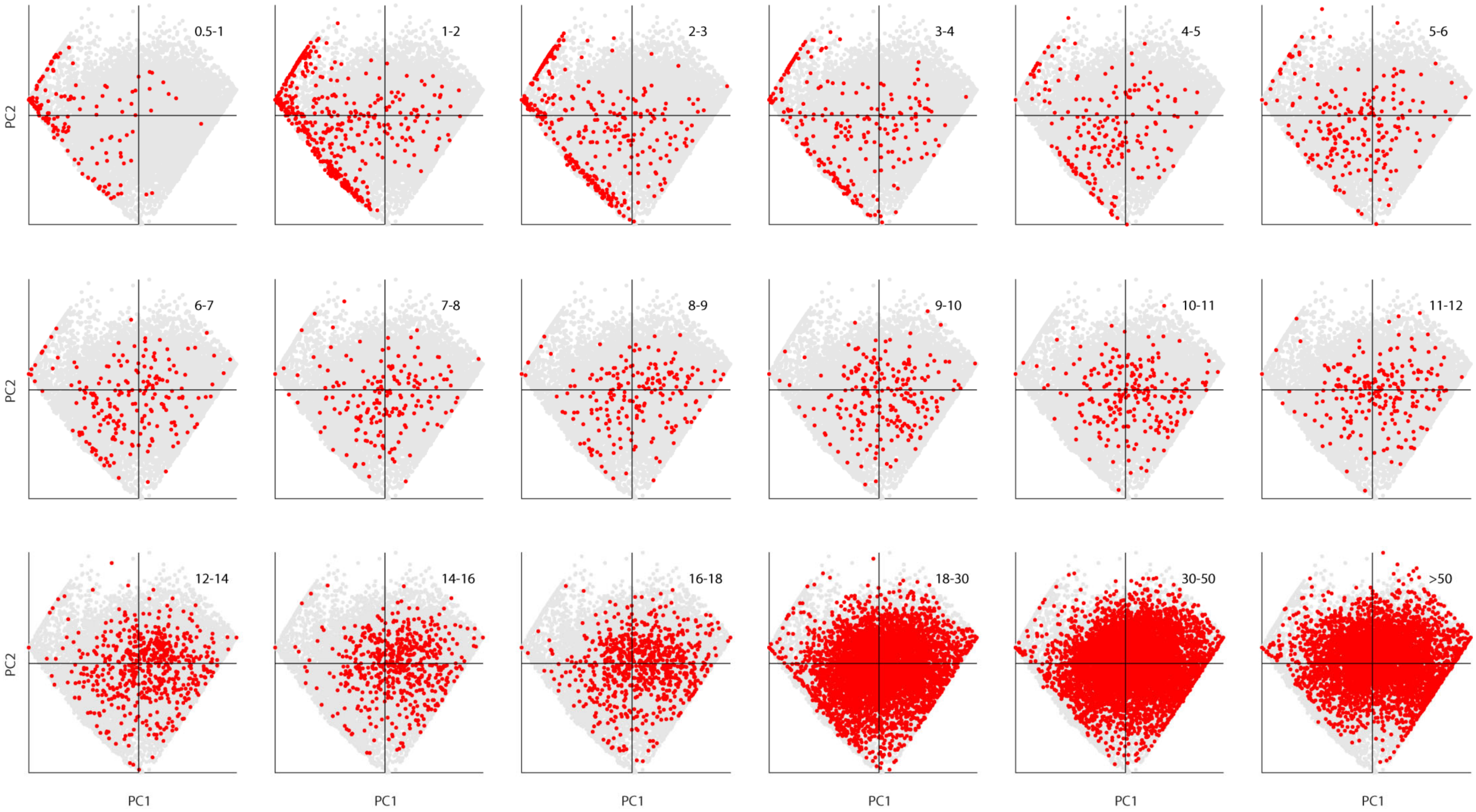


**Figure S11.** Scatter plots of individuals in principal components space, broken down by age group (shown in upper right of each panel). The grey dots in the background show all the data points. The red dots show the data points in each age class. Number of individuals (across all ages) shown here is $n$ = 24,402. This is the scatter-plot visualization of Figure 3 in the main paper.
